# Polygenic liability for major depression, schizophrenia and bipolar disorder and treatment outcome across major psychiatric disorders

**DOI:** 10.64898/2026.10.22.26366267

**Authors:** Alessandro Serretti, Daniel Souery, Siegfried Kasper, Gernot Fugger, Alexander Kautzky, Joseph Zohar, Stuart Montgomery, Panagiotis Ferentinos, Dan Rujescu, Raffaella Zanardi, Cristina Colombo, Francesco Benedetti, Diego Albani, Gianluigi Forloni, Stefano Barlati, Lisa Buson, Valentina Menesello, Paolo Martini, Rosana Carvalho Silva, Giulia Perusi, Gabriele Nibbio, Stefano Bignotti, PANDORA study group, Giovanni Battista Tura, Antonio Vita, Filippo Drago, Raffaele Ferri, Massimo Gennarelli, Alessandra Minelli, Eduard Vieta, Stefano Bonassi, Julien Mendlewicz

## Abstract

Polygenic scores for major depression, schizophrenia and bipolar disorder have been related to treatment outcome. We analysed 4,007 patients of European ancestry from 7 cohorts: three with major depression (STAR*D, GSRD, PANDORA), two with bipolar disorder (STEP-BD, Barcelona) and two with schizophrenia (CATIE, HSRRm). The three scores were computed with PRS-CS from the latest Psychiatric Genomics Consortium studies. The primary outcome was good response or good course, harmonized from each cohort’s instrument. Firth logistic regression within cohorts, adjusted for age, sex and ancestry components, was followed by random-effects pooling. A higher depression score was associated with lower odds of a good outcome (odds ratio per standard deviation 0.88, 95% confidence interval 0.79-0.99, p = .04; false discovery rate q = .11). The estimate remained below one in all 8 sensitivity analyses, including adjustment for baseline severity, and was negative in all 5 cohorts with a continuous improvement measure. The schizophrenia score pointed the same way (odds ratio 0.93, 95% confidence interval 0.85-1.01) with no heterogeneity. The bipolar score showed no pooled effect (odds ratio 0.97), and its direction differed by diagnosis. In pooled data spanning three diagnoses, higher polygenic liability to depression was nominally associated with a less favourable treatment outcome, although the association did not survive correction for multiple testing; schizophrenia liability showed a similar directional trend. These findings support further evaluation of disorder liability in multivariable models of treatment outcome.

## Introduction

Many patients with a major psychiatric disorder do not benefit enough from their first treatment. In the Sequenced Treatment Alternatives to Relieve Depression (STAR*D) study, fewer than half of the outpatients with major depressive disorder (MDD) responded to citalopram, and each further treatment step brought a smaller return [1,2]. About a third of patients with depression meet the current definition of treatment-resistant depression (TRD) [3]. In a nationwide cohort of adults with MDD, about one in four did not respond to the first antidepressant [4]. In schizophrenia, most patients in the Clinical Antipsychotic Trials of Intervention Effectiveness (CATIE) stopped their assigned antipsychotic before the end of the trial [5]. Between a fifth and a quarter of first-episode patients develop treatment-resistant schizophrenia [6], now defined by consensus criteria [7]. In bipolar disorder (BD), recurrence is frequent under guideline-based care [8], a substantial proportion of patients do not respond to lithium [9,10], and a definition of treatment-resistant bipolar depression has only recently been agreed [11]. Non-response is therefore common to all three disorders, and it carries much of their disability.

Clinical predictors explain part of this variability. In depression, the duration of the current episode, the number of previous episodes, hospitalizations and comorbid anxiety predict resistance [12–14]. Some plausible factors do not: moderate alcohol intake did not change antidepressant outcome in a prospective cohort [15]. Most of the variance in outcome remains unexplained. Genetic factors contribute to it. Common variants explain a small but significant part of the variance in antidepressant remission [16], and heritability estimates for treatment resistance vary with its definition [17–19]. An earlier estimate from pharmacogenetic trials was higher but imprecise [20]. Genome-wide association studies (GWAS) of response itself remain small, and none has produced a replicated locus [17,21,22].

A more powerful route uses the genetic liability to the disorders themselves. GWAS of schizophrenia, BD and depression now include hundreds of thousands of participants [23–25]. Polygenic scores (PGS) derived from them summarize each patient’s common-variant liability in a single number [26,27]. We refer to the scores for MDD, schizophrenia and BD as MDD-PGS, SCZ-PGS and BD-PGS. Bayesian methods such as PRS-CS use the full set of variants and predict better than scores built from a p-value threshold [28]. Whether these scores carry prognostic information beyond the diagnosis is an open question for clinical psychiatry [29,30]. Liability to depression is the most replicated polygenic signal for treatment outcome in MDD. In a meta-analysis of six European clinical samples, a higher MDD-PGS was nominally associated with non-remission and non-response to antidepressants [31], consistent in direction with the GENDEP and PGRN-AMPS cohorts [32]. The same direction appeared in the CAN-BIND-1 study [33] and in a Chinese sample treated with antidepressants [34]. In two ancestrally diverse biobanks, patients with TRD carried a higher MDD-PGS than treatment responders in every ancestry group [35]. The association is not limited to antidepressants. A higher MDD-PGS predicted less improvement after electroconvulsive therapy (ECT) in a large Swedish register sample [36] and a poorer response to lithium augmentation [37]. In BD, MDD-PGS was associated with poorer lithium response in the International Consortium on Lithium Genetics [38–40] and in the Mayo Clinic Bipolar Disorder Biobank [41]. A recent review estimated a 10% to 14% increase in the odds of non-remission per standard deviation of MDD-PGS [42]. Systematic reviews still describe the treatment literature as heterogeneous, with small samples and variable endpoints [43,44].

Schizophrenia liability has been linked to poorer outcome in all three disorders. In MDD, a higher SCZ-PGS was associated with non-response in the Group for the Study of Resistant Depression (GSRD) [45], and genetic risk for schizophrenia was negatively related to antidepressant response in the largest GWAS of response [16]. In first-episode psychosis, a higher SCZ-PGS predicted less improvement with antipsychotics in four cohorts [46]. A meta-analysis found a small and consistent association with treatment-resistant schizophrenia [47], and higher scores were related to clozapine use and to continuous hospitalization with resistance [48–50]. In BD, a higher SCZ-PGS predicted poorer lithium response [9]. These single-disorder findings have been summarized as a transdiagnostic effect of schizophrenia liability on treatment outcome [51]. Bipolar liability shows a different pattern. BD-PGS was unrelated to antidepressant response in MDD [52], whereas it predicted better response to ECT [36], to lithium augmentation [37] and to lithium in BD [41]. The direction of a BD-PGS effect may therefore depend on the disorder and on the treatment.

These observations fit the shared genetic architecture of the three disorders. Schizophrenia, BD and MDD share a large part of their common-variant liability [53–56]. Multivariate analyses place them on partly overlapping genetic factors [57,58], and pleiotropic loci shared by the three disorders converge on neurodevelopmental pathways [59]. A cross-trait analysis found shared polygenic susceptibility between treatment resistance in schizophrenia and lithium response in BD [60]. If liability shapes outcome through mechanisms shared across diagnoses, the association between a disorder PGS and outcome should appear in patients with other diagnoses as well.

The evidence has three limitations. First, most studies tested one score in one diagnosis, with different GWAS versions, scoring methods and outcome definitions, which prevents a direct comparison of effect sizes. Second, several studies analysed ancestrally mixed samples, although scores derived from European GWAS transfer poorly to other ancestries and are confounded by population structure [61]. Third, transdiagnostic studies have mostly examined symptom dimensions and clinical expression rather than treatment outcome [62,63]. Earlier genetic studies of these same cohorts were also limited to one diagnosis at a time: genome-wide predictors of long-term outcome in STEP-BD [64], schizophrenia risk loci and symptom dimensions in CATIE [65], shared psychopathological dimensions in the Italian and Spanish psychosis and bipolar samples [66], and schizophrenia liability and antidepressant non-response in GSRD [45,67]. Our group recently pooled trait PGS against treatment outcome in four cohorts across the three diagnoses; no association survived correction, and heterogeneity between diagnoses limited detection [68].

Four of the cohorts analysed here (STAR*D, GSRD, STEP-BD and CATIE) were also used in a pooled analysis from our group that is under review (Cantarella, submitted). That analysis had a different aim. It searched for clinical, sociodemographic and polygenic predictors of remission and treatment resistance, with outcomes defined on symptom severity and treatment failure. The present study examines the genetic liability to the three disorders and a broader outcome, response to treatment or course of illness, harmonized across seven cohorts, three of which were not in that analysis. The earlier analysis used scores from previous GWAS [69–71], and its pooled estimates for the three disorder scores were not significant. The latest GWAS are also more powerful than those used in the earlier analysis. The schizophrenia GWAS identified 287 loci, against 145 [23,71]. The bipolar score explains up to 9% of the variance in liability in European cohorts, a substantial improvement on the previous GWAS [24], and the depression score up to 5.8%, against 1.9% for the 2018 GWAS [25,69]. A single-cohort analysis of CATIE restricted to European ancestry found that a higher BD-PGS predicted less improvement and earlier discontinuation of antipsychotic treatment, a result we described as provisional until replicated (Serretti, submitted). Because the analyses use partially overlapping patients, they are not independent.

We therefore analysed individual participant data from seven cohorts covering MDD, BD and schizophrenia. The same three PGS, computed with one pipeline from the latest Psychiatric Genomics Consortium GWAS, were tested against a harmonized outcome of good response or good course in an ancestry-restricted primary analysis. We expected higher MDD-PGS and SCZ-PGS to be associated with lower odds of a good outcome across diagnoses, and we tested whether the BD-PGS association observed in CATIE extends to the other cohorts.

## Methods

### Samples

Seven cohorts contributed patient-level clinical and genome-wide data (Table 1). Three enrolled patients with MDD, two with BD and two with schizophrenia. Three are National Institute of Mental Health (NIMH) trials or effectiveness programmes and four are European clinical studies. Each parent study was approved by the ethics committee of its participating centres, as described in the original reports of STAR*D [72], GSRD [12], PANDORA [73], STEP-BD [74], the Barcelona cohort [75], CATIE [76] and HSRRm [66], and all participants gave written informed consent, including consent for genetic analyses. The present work is a secondary analysis of de-identified data. The n given for each cohort is the number of patients in the primary analysis; the parent studies were larger. STAR*D, GSRD, STEP-BD and CATIE were also used in earlier analyses from our group with other aims (see Introduction).

**Table 1.** Characteristics of the analysed sample, by cohort.

| Characteristic | CATIE | STAR*D | GSRD | STEP-BD | PANDORA | Barcelona | HSRRm |
| --- | --- | --- | --- | --- | --- | --- | --- |
| N analysed | 399 | 1255 | 1219 | 842 | 116 | 95 | 81 |
| Diagnosis | Schizophrenia | MDD | MDD (treatment-resistant sample) | Bipolar disorder | MDD | Bipolar disorder | Schizophrenia |
| Treatment | Quetiapine 94;<br>Risperidone 92;<br>Olanzapine 90;<br>Perphenazine 75;<br>Ziprasidone 48 | citalopram<br>(Level 1) | naturalistic antidepressant treatment | naturalistic, guideline-based care | Duloxetine 28;<br>Sertraline 23;<br>Venlafaxine 14;<br>Escitalopram 12;<br>Paroxetine 11;<br>Fluoxetine 6;<br>Bupropion 6;<br>Vortioxetine 4;<br>Fluvoxamine 4;<br>Citalopram 2;<br>Amitriptyline 2;<br>Nortriptyline 2;<br>Mirtazapine 1;<br>Amisulpride 1 | lithium 85;<br>valproate 10 | naturalistic antipsychotic treatment |
| Age, years, mean (SD) | 41.3 (11.4) | 42.9 (13.6) | 51.6 (13.9) | 41.3 (12.5) | 48.1 (14.2) | 47.4 (13.0) | 50.3 (13.6) |
| Female, n (%) | 91 (22.8%) | 724 (57.7%) | 788 (64.6%) | 465 (55.2%) | 81 (69.8%) | 41 (43.2%) | 43 (53.1%) |
| Education, years, mean (SD) | 12.4 (2.2); n=398 | 14.2 (2.9); n=1253 | 11.9 (4.2); n=1211 | 14.8 (2.1); n=796 | 12.8 (3.6) | 11.6 (3.7); n=83 | 10.7 (3.3); n=77 |
| Employed, n (%) | 66 (16.7%); n=396 | 758 (60.4%) | 499 (48.1%); n=1038 | 348 (43.9%); n=793 | 80 (69.0%) | 39 (43.8%); n=89 | 7 (10.1%); n=69 |
| Married or cohabiting, n (%) | 46 (11.5%) | 557 (44.4%) | 597 (49.0%) | 310 (38.9%); n=796 | 48 (41.7%); n=115 | 31 (34.4%); n=90 | n.a. |
| Age at onset, years, mean (SD) | 24.1 (9.0); n=384 | 25.1 (14.5); n=1246 | 36.7 (15.0); n=1164 | 22.8 (10.2); n=800 | 33.8 (14.9) | 27.3 (10.1); n=86 | 27.9 (19.9); n=76 |
| Lifetime suicide attempt, n (%) | n.a. | 175 (14.0%); n=1254 | n.a. | 339 (41.4%); n=818 | 14 (12.1%) | 28 (32.2%); n=87 | n.a. |
| Family history of psychiatric disorder, n (%) | n.a. | 775 (62.4%); n=1242 | 464 (38.1%); n=1217 | n.a. | 67 (57.8%) | 66 (79.5%); n=83 | 43 (53.8%); n=80 |
| Comorbid anxiety disorder, n (%) | 74 (18.5%) | 141 (15.0%); n=942 | 223 (18.3%); n=1218 | 441 (57.0%); n=774 | 33 (28.4%) | n.a. | n.a. |
| Substance abuse or dependence, n (%) | 129 (32.3%) | 80 (26.6%); n=301 | 53 (4.4%); n=1217 | 427 (55.1%); n=775 | 5 (4.3%) | 15 (17.4%); n=86 | 25 (30.9%) |
| Baseline severity, mean (SD) | 72.9 (17.5) [PANSS total (30- | 15.1 (4.3) [QIDS-SR16 | 35.0 (7.6) [MADRS (0-60), retrospective rating at | 16.2 (11.4); n=782 [MADRS (0-60) at | 17.3 (3.3) [HAM-D total at T0] | n.a. | n.a. |
|  | 210)] | (0-27)] | the start of the treatment] | study entry] |  |  |  |
| Primary outcome definition | PANSS reduction $\geq 20\%$ (Phase 1/1A) | QIDS-SR response ($\geq 50\%$) | MADRS responder (vs non-responder or resistant) | Percent of days well above the median | HAM-D response at week 12 | Good response to lithium (valproate if no lithium) | OPCRIT item 90: course without chronic deterioration |
| Good response or course, n (%) | 161 (40.4%) | 662 (52.7%) | 338 (27.7%) | 420 (49.9%) | 75 (64.7%) | 80 (84.2%) | 37 (45.7%) |
| Improvement, %, mean (SD) | 4.8 (51.3) | 46.7 (34.2) | 30.0 (32.3) | 52.6 (31.3) | 58.0 (25.3) | n.a. | n.a. |
| SCZ-PGS weight-file variants scored, % | 99.6 | 99.4 | 100.0; 98.6 | 94.2 | 99.6 | 67.7 | 99.7 |

GSRD combines two genotyping batches, analysed in one model with a batch indicator and batch-specific principal components (PGS standardized within batch; SCZ-PGS coverage shown per batch). Patients analysed in the primary model (ancestry-restricted, complete data for outcome, PGS, age, sex and principal components). n shown when a variable is incomplete; n.a., not recorded in that cohort. Education in GSRD and STEP-BD converted from attainment level to typical years. Family history: mood disorder in a relative (STAR\*D, GSRD), concordant or discordant psychiatric history (PANDORA), any psychiatric illness (Barcelona), OPCRIT items 13-14 (HSRRm). Substance use: SCID past 5 years (CATIE), PDSQ alcohol or cannabis (STAR\*D), alcohol only (PANDORA). Improvement: QIDS-SR (STAR\*D), MADRS (GSRD), HAM-D (PANDORA), baseline-corrected PANSS (CATIE), percent of days well (STEP-BD). Baseline severity is shown on each cohort's own instrument. PGS, polygenic score; SCZ, schizophrenia.

#### STAR*D (n = 1,255)

The Sequenced Treatment Alternatives to Relieve Depression study was a multisite, prospective, multistep trial in outpatients aged 18 to 75 years with nonpsychotic MDD, enrolled from primary and specialty care in the United States [72]. All participants started on citalopram at level 1, the treatment step analysed here [1]. Depressive symptoms were rated with the 16-item Quick Inventory of Depressive Symptomatology, self-report (QIDS-SR16) [77].

#### GSRD (n = 1,219)

The Group for the Study of Resistant Depression is a European multicentre project on treatment resistance in depression, recruiting since 2000 in mental health centres in several countries [12,78]. Adults with a major depressive episode are treated naturalistically with antidepressants at adequate dose for at least four weeks. The Montgomery-Åsberg Depression Rating Scale (MADRS) [79] is rated at inclusion and, from interview and medical records, for the beginning of the current episode. Patients are classified as responders, non-responders or treatment resistant, the last defined as non-response to at least two adequate antidepressant trials [67,80]. Patients of the earlier recruitment waves [45] and of the fourth wave [14] were genotyped in two separate batches.

#### PANDORA (n = 116)

PANDORA is an Italian randomized controlled trial of a combinatorial pharmacogenomic test in MDD [73]. Patients starting a new antidepressant, because the previous one had failed or was not tolerated, were rated with the Hamilton Depression Rating Scale (HAM-D) [81] at baseline and at weeks 4, 8 and 12 [82].

#### STEP-BD (n = 842)

The Systematic Treatment Enhancement Program for Bipolar Disorder was an NIMH effectiveness programme in patients with BD of any subtype, followed prospectively under guideline-based care [74]. Clinical status was recorded at every visit, allowing the proportion of follow-up time spent well to be computed [8].

#### Barcelona bipolar cohort (n = 95)

Outpatients of the BD Unit of the Hospital Clinic, University of Barcelona, with DSM-IV-TR bipolar I or II disorder confirmed with the Structured Clinical Interview for DSM-IV, were enrolled in a naturalistic cohort [66,75]. Long-term response to lithium, and to valproate in patients without a lithium rating, was rated by the treating psychiatrists as good, partial or poor.

#### CATIE (n = 399)

The Clinical Antipsychotic Trials of Intervention Effectiveness compared perphenazine with olanzapine, quetiapine, risperidone and ziprasidone in patients with chronic schizophrenia at 57 sites in the United States, with double-blind treatment for up to 18 months [5,76]. We analysed Phase 1/1A, in which all participants were randomized to one of the study drugs. Symptoms were rated with the Positive and Negative Syndrome Scale (PANSS) [83].

#### HSRRm (n = 81)

Inpatients with a DSM-IV-TR schizophrenia-spectrum diagnosis were recruited at two facilities in Rome, the San Filippo Neri Hospital and the San Raffaele Villa dei Fiori residential unit [66]. Lifetime diagnosis and course were rated with the Operational Criteria Checklist for Psychotic Illness (OPCRIT) [84].

### Outcomes

The primary outcome was a binary indicator of good response or good course, built from each cohort’s own instrument before any genetic association was computed (S4 Table). Five cohorts measured response to a treatment. In STAR*D it was a reduction of at least 50% in QIDS-SR16 at the end of level 1 [1]. In GSRD, response was defined as reduction of at least 50% in MADRS after at least four weeks of adequate treatment; non-responders and treatment-resistant patients were combined as the comparison group [67]. In PANDORA it was a reduction of at least 50% in HAM-D at week 12. In CATIE response was a reduction of at least 20% in PANSS total at the last Phase 1/1A visit, computed after subtraction of the 30-point floor of the scale [85,86]. In the Barcelona cohort it was good long-term response to lithium, or to valproate when lithium had not been rated, against partial or poor response. Two cohorts measured illness course. In STEP-BD it was a proportion of follow-up days spent well above the median of the whole STEP-BD registry, and in HSRRm it was any OPCRIT course of disorder other than continuous chronic illness with deterioration (item 90, codes 1 to 4 against code 5).

The secondary outcome was the percentage improvement, standardized within cohort, in the five cohorts with a continuous measure: QIDS-SR16 in STAR*D, MADRS in GSRD, HAM-D in PANDORA, PANSS in CATIE (with the same floor correction) and the percentage of days well in STEP-BD. A higher value means a better outcome in every cohort.

### Genotyping, imputation and quality control

Each cohort was genotyped and quality controlled by its own group. CATIE was genotyped on the Illumina HumanHap550 array [87], STAR*D on the Affymetrix 500K and 5.0 arrays [88], and STEP-BD on the Affymetrix 500K array [89]. GSRD was genotyped on the Illumina Infinium PsychArray [90], PANDORA on the Illumina PsychArray-24 and Global Screening Array-24, and the Barcelona and HSRRm cohorts on the Illumina PsychArray, processed with the Psychiatric Genomics Consortium quality control and imputation pipeline [66,91]. Standard filters removed samples and variants with high missingness, sex discrepancy or abnormal heterozygosity, and genotypes were imputed to the Haplotype Reference Consortium panel [67,92]. Hardy-Weinberg equilibrium was not used as a common filter, because all samples consisted of patients, in whom departures from equilibrium are expected [66]. PANDORA and HSRRm were imputed for this study on the Michigan Imputation Server with the Haplotype Reference Consortium r1.1 panel [93]. Variants with imputation R² below 0.3 or minor allele frequency below 0.01 were discarded, and scores were computed from dosages.

### Polygenic scores

Scores for schizophrenia, BD and MDD were computed with PRS-CS [28] from the European-ancestry summary statistics of the latest Psychiatric Genomics Consortium GWAS: schizophrenia wave 3 [23], BD wave 4 [24] and MDD 2025 without the 23andMe contribution [25]. PRS-CS inferred posterior effect sizes with the 1000 Genomes European linkage-disequilibrium reference, default shrinkage hyperparameters and a fixed global shrinkage parameter of 0.01. The same three weight files were applied in every cohort with PLINK [94]. They contained 1,107,744 variants for SCZ-PGS, 1,089,999 for BD-PGS and 1,049,606 for MDD-PGS. The proportion of SCZ-PGS weight-file variants scored was at least 94.2% in six cohorts and 67.7% in the Barcelona cohort (Table 1). Scores were standardized within cohort and within the analysis set, and within genotyping batch in GSRD, so that every estimate is per standard deviation (SD) of the cohort’s own score distribution.

Six cohorts contributed to the discovery GWAS of their own disorder: CATIE and HSRRm to schizophrenia, STEP-BD and the Barcelona cohort to BD, and STAR*D and the earlier GSRD batch to MDD. PANDORA and the fourth GSRD batch did not. Sample overlap inflates the association of a score with the discovery phenotype, here the diagnosis, which is constant within each cohort. Within an all-patient cohort it adds variance to the own-disorder score and, if anything, biases its per-SD association with outcome toward the null. We tested its influence with an overlap-free sensitivity analysis that pooled each score over the cohorts, or genotyping batches, outside its discovery GWAS [26].

### Ancestry and relatedness

The primary analysis was restricted to patients of European genetic ancestry, the ancestry of the discovery GWAS [61]. Ancestry was defined statistically from genotype data to match the discovery samples of the scores, and the label does not describe social or cultural identity. Sex was taken from each cohort’s clinical record and coded as female or male. In CATIE, we used the European-ancestry set defined from genotype data by the NIMH repository, as in the pilot analysis (Serretti, unpublished data). In the other cohorts, principal components were computed on linkage-disequilibrium-pruned genotypes of unrelated samples with PLINK 2, and all samples were projected onto them. A patient was kept if they lay within 6 SD of the reference-group mean on each of the first 10 components, the outlier criterion of EIGENSOFT [95]. The reference group was the self-reported White non-Hispanic participants in STAR*D and STEP-BD, and all genotyped samples elsewhere. Principal components were then recomputed within the retained samples of each cohort, and the first five entered the primary models. Relatedness was estimated with the KING-robust kinship coefficient within each cohort and between the two GSRD batches [96]. When a genotype duplicate (kinship above 0.354) was linked to two different clinical records, both records were excluded, because the assignment of DNA to record could not be verified. For related pairs (kinship above 0.0884), one member was excluded.

### Covariates and descriptive variables

All models were adjusted for age, sex and the first five within-cohort principal components. Socio-demographic and clinical characteristics were harmonized for description only: education in years, employment, partnership, age at onset, lifetime suicide attempt, family history of psychiatric disorder, comorbid anxiety disorder, comorbid substance use disorder and baseline severity (Table 1). Education recorded as an attainment level in GSRD and STEP-BD was converted to typical years. S4 Table gives the source variable, the coding rule and the coverage of every harmonized variable in every cohort.

### Statistical analysis

We used a two-stage individual participant data design [97]. In stage one, each cohort was analysed separately, with one model per score: Firth penalized logistic regression of the primary outcome on the standardized score, age, sex and five principal components, which gives a log odds ratio per SD, reported as an odds ratio (OR). Firth estimation [98] was used in every cohort, because three cohorts had fewer than 120 patients and few events in one outcome class, a setting in which maximum likelihood estimates are biased. In the 12 fits with at least 50 patients in each outcome class, Firth and maximum likelihood estimates differed by at most 0.008 on the log-odds scale. GSRD entered as one cohort, with a batch indicator and batch-specific principal components. The secondary outcome was analysed with ordinary least squares and the same covariates. In stage two, cohort estimates were pooled with random-effects models, with restricted maximum likelihood estimation of the between-cohort variance and Hartung-Knapp-Sidik-Jonkman confidence intervals [99]; p-values came from a t distribution with k-1 degrees of freedom. Heterogeneity was summarized with I² and Cochran’s Q. All intervals are 95% confidence intervals (CI). Patient records were not pooled into a single model, because cohort, diagnosis and treatment are confounded by design.

Benjamini-Hochberg false discovery rate (FDR) correction was applied to the three primary tests, one per score. The secondary outcome, the sensitivity and subgroup analyses and the all-ancestry arm were exploratory and are reported without correction. Eight sensitivity analyses repeated the primary model with 10 principal components; with the three scores in one model; in the five cohorts with a treatment-response outcome; with GSRD responders compared only with treatment-resistant patients; with the CATIE outcome defined as a drop of at least one point in Clinical Global Impression severity; without CATIE, the pilot cohort; without the cohorts in each score’s discovery GWAS; and with adjustment for standardized baseline severity in the five cohorts with a pre-treatment total, compared with the unadjusted model in the same patients. We also left out one cohort at a time. Pooled estimates were also computed within diagnosis, with a between-subgroup Q test. An exploratory arm repeated the primary model in all genotyped patients regardless of ancestry, with full-sample principal components.

Nominal significance was defined as a two-sided p below 0.05, and a false discovery rate q below 0.05 was required for an association to be considered significant after correction. Patients without the primary outcome or a covariate were excluded from the analysis. No sample size calculation was performed, because this is a secondary analysis of all patients with genetic and outcome data; the consequences for power are discussed under Strengths and limitations. Analyses used Python 3.11.12 (pandas 2.3.3, NumPy 2.4.2, statsmodels 0.14.6, SciPy 1.17.1), with the Firth estimator implemented for this study, and PLINK v1.90b7.2 and PLINK v2.0.0-a.6.12 for genotype processing. Reporting follows the STREGA extension of the STROBE statement for genetic association studies [100] and the Polygenic Risk Score Reporting Standards [101]; the completed STREGA checklist is S1 Checklist.

### Use of generative AI

We used Anthropic Claude Code (Claude Sonnet) and OpenAI Codex during the study. They helped to brainstorm the analysis plan, to write and refactor the Python scripts for data harmonization, models, tables and figures, and to draft and edit the manuscript text. They did not generate or alter any data. All numbers in the text were inserted automatically from the saved analysis outputs, and a script checked that no statistic was typed by hand. The Firth estimator was compared with maximum likelihood estimation in the fits with enough events in both outcome classes (see Statistical analysis). References were retrieved and verified in PubMed. The authors read and edited all text and code outputs and take full responsibility for the content of the article.

## Results

### Sample

Of 4,452 patients with clinical and genetic data, 17 were excluded at genetic quality control (duplicates and relatives) and 126 lacked the primary outcome (Fig 1). Of the remaining 4,309, 296 fell outside the European ancestry set, most of them in STAR*D (204) and CATIE (71), and 6 lacked a covariate. The primary analysis included 4,007 patients: 2,590 with MDD, 937 with BD and 480 with schizophrenia. Mean age was 45.5 years (SD 14.0) and 2,233 patients (55.7%) were women. Overall, 1,773 of 4,007 patients (44.2%) had a good outcome, from 338 of 1,219 (27.7%) in GSRD, a sample enriched for treatment resistance, to 80 of 95 (84.2%) in the Barcelona cohort (Table 1). The three largest cohorts, STAR*D, GSRD and STEP-BD, contributed 3,316 of 4,007 patients (82.8%).

**Fig 1.**
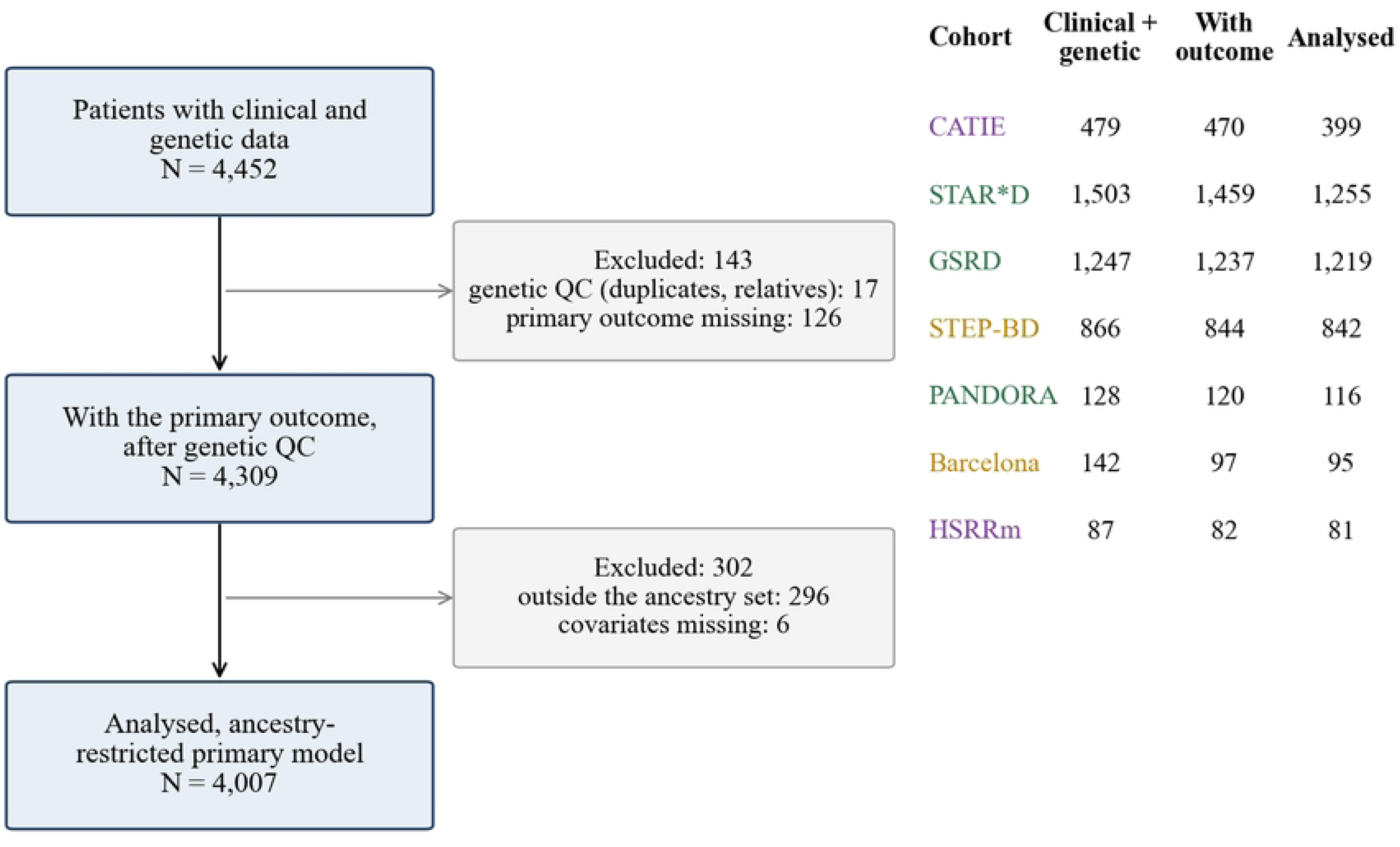
Patient flow. Left, from the patients with both clinical and genetic data to the sample analysed in the primary, ancestry-restricted model, with the number excluded at each step and the reason. Right, the same counts per cohort. QC, quality control.

### Primary analysis

Table 2 and Fig 2 give the cohort and pooled estimates, and Table 3 summarizes the pooled results. A higher MDD-PGS was associated with lower odds of a good outcome (OR per SD 0.88, CI 0.79-0.99, p = .04), about 12% lower odds per SD, with low heterogeneity (I² = 18%). The association did not survive FDR correction across the three primary tests (q = .11). The cohort estimate was below one in 4 of 7 cohorts, including the three largest, and was nominally significant in GSRD (OR 0.81, CI 0.71-0.93) and STEP-BD (OR 0.80, CI 0.69-0.92). The three cohorts with an estimate above one, CATIE, Barcelona and HSRRm, had wide intervals that included the pooled estimate.

**Table 2.** Cohort-level and pooled associations between each polygenic score and good response or good course.

| Cohort | N | Good response/course,<br>n (%) | SCZ-PGS OR (95% CI) | SCZ-PGS p | BD-PGS OR (95% CI) | BD-PGS<br>p | MDD-PGS OR (95%<br>CI) | MDD-PGS p |
| --- | --- | --- | --- | --- | --- | --- | --- | --- |
| CATIE | 399 | 161 (40.4%) | 0.95 (0.77-1.17) | .62 | 0.85 (0.68-1.06) | .14 | 1.01 (0.83-1.24) | .91 |
| STAR*D | 1255 | 662 (52.7%) | 0.96 (0.85-1.09) | .54 | 0.94 (0.83-1.06) | .32 | 0.92 (0.82-1.04) | .17 |
| GSRD | 1219 | 338 (27.7%) | 0.84 (0.73-0.97) | .01 | 0.90 (0.78-1.03) | .14 | 0.81 (0.71-0.93) | .002 |
| STEP-BD | 842 | 420 (49.9%) | 0.98 (0.85-1.12) | .74 | 1.20 (1.04-1.38) | .01 | 0.80 (0.69-0.92) | .001 |
| PANDORA | 116 | 75 (64.7%) | 1.16 (0.72-1.85) | .54 | 1.03 (0.68-1.58) | .87 | 0.93 (0.60-1.44) | .75 |
| Barcelona | 95 | 80 (84.2%) | 0.73 (0.38-1.40) | .34 | 0.90 (0.48-1.68) | .73 | 1.10 (0.58-2.07) | .78 |
| HSRRm | 81 | 37 (45.7%) | 0.67 (0.40-1.13) | .13 | 0.97 (0.60-1.56) | .90 | 1.11 (0.68-1.81) | .67 |
| Pooled (random<br>effects) | 4007 |  | 0.93 (0.85-1.01) | .08 | 0.97 (0.84-1.13) | .68 | 0.88 (0.79-0.99) | .04 |
| Heterogeneity |  |  | I<sup>2</sup> = 0%, tau<sup>2</sup> = 0.000 | Q p = .46 | I<sup>2</sup> = 48%, tau<sup>2</sup> = 0.012 | Q p = .07 | I<sup>2</sup> = 18%, tau<sup>2</sup> = 0.004 | Q p = .29 |

Odds ratio of good response or course per SD of each PGS, one model per PGS, Firth penalized logistic regression. Adjusted for age, sex and the first 5 ancestry principal components recomputed within each cohort's ancestry-restricted sample. Pooled: random effects, REML tau<sup>2</sup>, Hartung-Knapp-Sidik-Jonkman confidence interval and p (t, k-1 df). OR < 1: higher PGS, lower odds of good outcome. OR, odds ratio.

**Fig 2.**
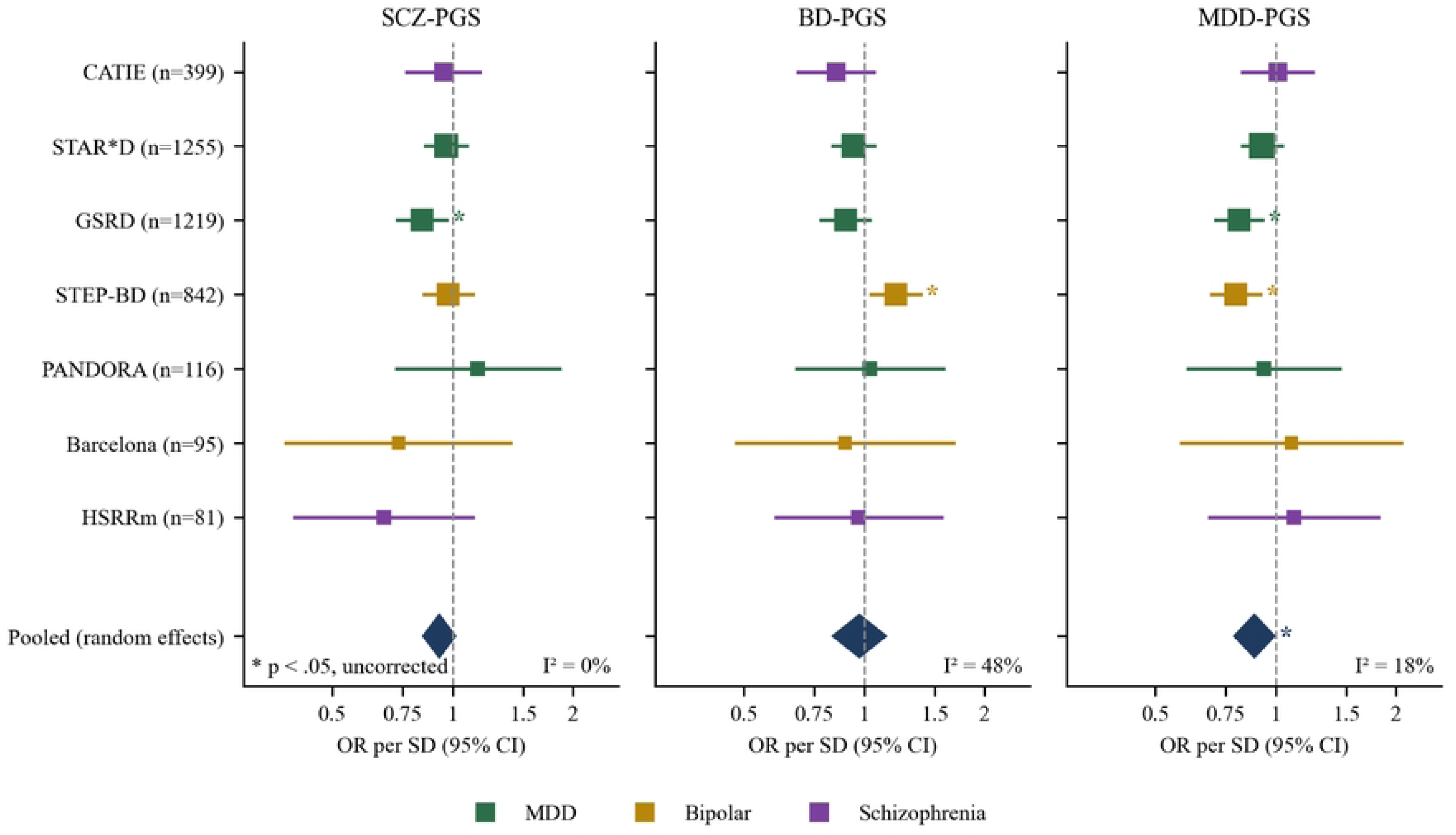
Association between each polygenic score and good outcome, by cohort and pooled. Squares are cohort odds ratios per SD of score with 95% confidence intervals from Firth logistic regression adjusted for age, sex and five ancestry principal components; square size is proportional to the inverse-variance weight and colour marks the diagnosis. Diamonds are pooled random-effects odds ratios (restricted maximum likelihood, Hartung-Knapp-Sidik-Jonkman interval). An odds ratio below 1 means that a higher score is associated with lower odds of a good outcome. Arrowheads mark intervals that extend beyond the axis. An asterisk marks nominal significance (p < .05, uncorrected). PGS, polygenic score; SCZ, schizophrenia; BD, bipolar disorder; MDD, major depressive disorder.

**Table 3.** Pooled estimates of the primary analysis.

| PGS | Response:<br>cohorts (k) | Response:<br>N | Response OR (95%<br>CI) | Response<br>p | Response<br>FDR q | Response<br>I<sup>2</sup> | Improvement beta (95%<br>CI) | Improvement<br>t p | Improvement:<br>cohorts (k) | Improvement<br>t I<sup>2</sup> |
| --- | --- | --- | --- | --- | --- | --- | --- | --- | --- | --- |
| SCZ-PGS | 7 | 4007 | 0.93 (0.85-1.01) | .08 | .12 | 0% | -0.05 (-0.10 to 0.01) | .09 | 5 | 21% |
| BD-PGS | 7 | 4007 | 0.97 (0.84-1.13) | .68 | .68 | 48% | -0.00 (-0.10 to 0.09) | .98 | 5 | 68% |
| MDD-PGS | 7 | 4007 | 0.88 (0.79-0.99) | .04 | .11 | 18% | -0.08 (-0.16 to -0.00) | .05 | 5 | 55% |

Primary analysis, ancestry-restricted. Response: odds ratio per SD of PGS for good response or course, all 7 cohorts. FDR q: Benjamini-Hochberg false discovery rate over the three primary tests. Improvement: standardized percentage improvement per SD of PGS (OLS, higher = better), five cohorts with a continuous measure; reported uncorrected. Adjusted for age, sex and the first 5 ancestry principal components recomputed within each cohort's ancestry-restricted sample.

The SCZ-PGS estimate had the same direction (OR 0.93, CI 0.85-1.01, p = .08, q = .12). The estimate was below one in 6 of 7 cohorts, with no heterogeneity (I² = 0%), and was nominally significant in GSRD (OR 0.84, CI 0.73-0.97). BD-PGS showed no pooled association (OR 0.97, CI 0.84-1.13, p = .68) and moderate heterogeneity (I² = 48%). Its estimates ranged from 0.85 in CATIE to 1.20 in STEP-BD, where a higher BD-PGS was associated with a better course (CI 1.04-1.38).

### Secondary outcome

On standardized percentage improvement (Table 3, S5 Table), a higher MDD-PGS was associated with less improvement (β per SD -0.08, CI -0.16 to -0.002, p = .05). The estimate was negative in all 5 cohorts with a continuous measure and nominally significant in STAR*D and STEP-BD. SCZ-PGS showed the same direction (β -0.05, p = .09), and BD-PGS showed no association (p = .98).

### Sensitivity analyses

The MDD-PGS estimate was stable (S2 Table, Fig 3). It was below one in all 8 sensitivity analyses of the primary outcome and nominally significant in 5: with 10 principal components (OR 0.89, p = .04), with GSRD responders compared only with treatment-resistant patients (OR 0.89, p = .04), with the CATIE outcome defined on Clinical Global Impression severity (OR 0.86, p = .009), without CATIE (OR 0.86, CI 0.77-0.97, p = .02), and after adjustment for baseline severity (OR 0.89, p = .04, against 0.87 without adjustment in the same patients). Leaving out one cohort at a time gave ORs in the range 0.86-0.91. The overlap-free estimate, from 6 cohorts or batches outside the MDD discovery GWAS, had the same size as the primary estimate (OR 0.88, CI 0.73-1.07) with a wider interval (p = .16). The model with the three scores together gave OR 0.90 (p = .14), and the five treatment-response cohorts OR 0.90 (p = .13). In the exploratory all-ancestry arm (4,303 patients; S1 Table and S3 Table), the estimate was OR 0.89 (CI 0.79-0.99, p = .03). On the secondary outcome, adjustment for baseline severity left the MDD-PGS estimate similar (β -0.07, against -0.08 without adjustment in the same patients), reduced heterogeneity (I² = 0%, against 53%) and narrowed the interval (CI -0.11 to -0.02, p = .01).

**Fig 3.**
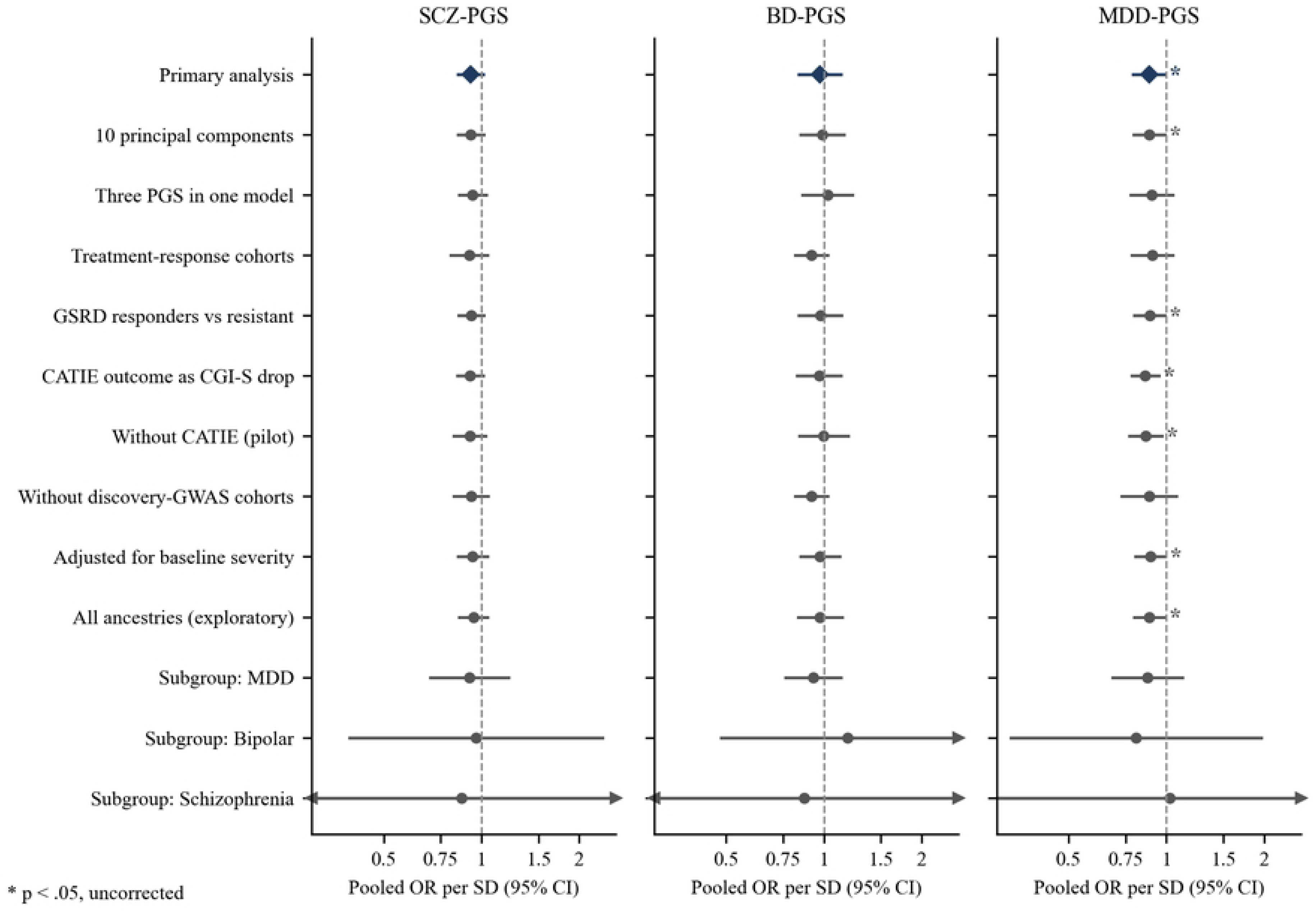
Pooled odds ratios across the primary, sensitivity and subgroup analyses. Pooled odds ratios per SD of each polygenic score across the primary analysis (diamond), the sensitivity analyses, the exploratory all-ancestry arm and the diagnosis subgroups (circles), with 95% confidence intervals. Arrowheads mark intervals that extend beyond the axis. An asterisk marks nominal significance (p < .05, uncorrected). GSRD, Group for the Study of Resistant Depression; CATIE, Clinical Antipsychotic Trials of Intervention Effectiveness; CGI-S, Clinical Global Impression severity.

The SCZ-PGS estimate was below one in all 8 sensitivity analyses, and leaving out one cohort at a time gave ORs in the range 0.91-0.96. BD-PGS remained null in every sensitivity analysis; without CATIE its pooled OR was 1.00 (p = .98).

### Diagnosis subgroups

MDD-PGS and SCZ-PGS estimates did not differ between MDD, BD and schizophrenia (between-subgroup p = .13 and p = .78). BD-PGS estimates differed by diagnosis (p = .007): OR 1.18 in BD, 0.93 in MDD and 0.87 in schizophrenia, with the bipolar estimate driven by STEP-BD (S2 Table).

## Discussion

To our knowledge, this is the first individual participant data analysis to test the newest disorder-liability polygenic scores against treatment outcome across MDD, BD and schizophrenia with a single scoring pipeline. Across seven cohorts, a higher polygenic liability to major depression was associated with a less favourable outcome, about 12% lower odds of good response or good course per SD. The association was small, homogeneous and stable. Its direction remained consistent across all sensitivity analyses. It remained nominally significant after adjustment for baseline severity, without the pilot cohort and in all ancestries, and on the continuous improvement outcome all 5 cohorts pointed the same way. It did not survive correction across the three primary tests and should therefore be considered suggestive rather than definitive; nevertheless, its consistency across sensitivity analyses and the absence of significant between-diagnosis heterogeneity support further investigation of MDD polygenic liability as a potential transdiagnostic prognostic marker. Schizophrenia liability pointed the same way, with a smaller estimate and no heterogeneity. Bipolar liability had no overall effect, and the association with poorer outcome that we had observed in CATIE did not extend to the other cohorts. Fig 4 summarizes the sample, the pooled estimates and the main numbers for the depression score.

**Fig 4.**
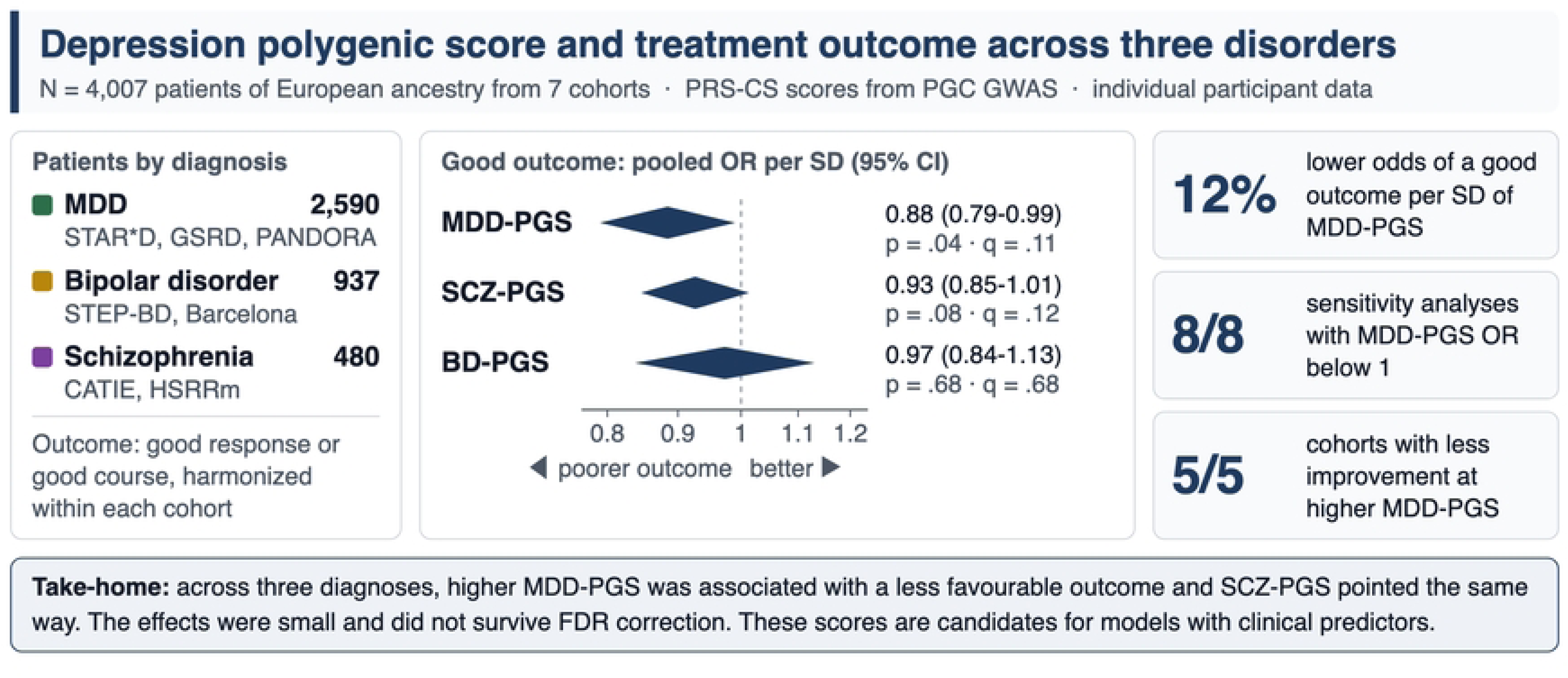
Graphical abstract of the study and its main findings. Left, patients by diagnosis and cohort. Centre, pooled odds ratios per SD of each polygenic score for good response or good course in the primary analysis, with 95% confidence intervals, p-values and false discovery rate q-values. Right, the percentage lower odds of a good outcome per SD of MDD-PGS, the number of sensitivity analyses in which the MDD-PGS odds ratio was below 1, and the number of cohorts with less improvement at a higher MDD-PGS. FDR, false discovery rate.

The size of the MDD-PGS association matches earlier estimates obtained with other designs and treatments. Our estimate corresponds to about 13% higher odds of a poor outcome per SD. A recent review estimated 10% to 14% higher odds of non-remission per SD [42], and an OR of 0.89 per SD was reported for improvement after ECT [36]. Treatment resistance defined in biobanks from prescriptions or ECT also carried a higher polygenic burden for MDD and related disorders [18,19]. The present analysis brings these single-disorder observations into one pooled model that spans three diagnoses, several drug classes and two measures of illness course. The data are compatible with a prognostic association that may not be specific to a particular treatment. Cohort estimates were below one under citalopram (STAR*D), under naturalistic antidepressant treatment (GSRD, PANDORA) and for days well under guideline-based care (STEP-BD). In CATIE, Barcelona and HSRRm, treated with antipsychotics or mood stabilizers, the estimates were at or slightly above one, with wide intervals that included the pooled estimate. Heterogeneity was low, and the estimates did not differ significantly between diagnoses. Baseline severity did not explain the association: adjustment left the odds ratio almost unchanged and reduced the heterogeneity of the association with percentage improvement. MDD-PGS has been associated with symptom burden and course in depression [44], and it is genetically correlated with neuroticism [69,102]. Polygenic scores for neuroticism, insomnia, as well as related traits, were associated with TRD in two large biobanks [103,104]. Sleep and circadian dysregulation may represent an additional pathway linking polygenic liability to clinical outcome, since sleep disturbance has been associated with persistence, relapse and poorer clinical course across mood and psychotic disorders. A higher MDD-PGS may therefore mark a disposition to persistent negative affect and recurrence that limits the benefit of any treatment. This interpretation fits the observation that MDD-PGS predicted poorer outcome after ECT [36], a treatment with a mechanism distinct from that of antidepressant drugs.

Schizophrenia liability gave a smaller signal with the same direction. Earlier studies found this direction with antidepressants, antipsychotics and lithium [9,16,46,47], although not in every sample [105]. One of them analysed the first GSRD batch, which overlaps with the present sample [45]. In our data the SCZ-PGS estimate was below one in 6 of 7 cohorts, with I² = 0%, so the pooled estimate reflects a small shift shared by most cohorts. Two mechanisms are plausible. The first involves cognitive function. Treatment resistance in schizophrenia shares genetic correlations with cognitive traits [48]. In our earlier multi-cohort analysis, a PGS indexing worse executive function was nominally associated with non-remission [68], and liability to lower educational attainment predicted relapse after a first psychotic episode [106]. The second is treatment adherence. SCZ-PGS was associated with poorer adherence to lithium [39], and adherence is lowest in patients with schizophrenia and BD in clinical cohorts [107]. The ECT literature adds a caveat. A higher SCZ-PGS predicted better response to ECT in one study [108], so the direction may depend on the treatment.

Bipolar liability behaved differently. The pooled estimate was null and heterogeneous, and the estimates differed by diagnosis. In STEP-BD a higher BD-PGS was associated with more days well, while in the other cohorts the estimates were close to one or below. Earlier studies also found a BD-PGS direction that depends on diagnosis or treatment [36,37,41,52]. In BD, a higher BD-PGS may mark a more typical and episodic illness, the profile that responds best to mood stabilizers [109]. Clinical features also modify treatment response in BD, for example comorbid borderline personality disorder [110], and such features may dilute a genetic signal in naturalistic samples. The pooled BD-PGS estimate without CATIE was null, so the association observed in CATIE did not extend to the other cohorts. The only other schizophrenia cohort, HSRRm, was small, and its estimate was close to one (OR 0.97). The pilot finding should be considered specific to CATIE until a larger schizophrenia cohort replicates it.

Our data suggest that liability to depression, and possibly to schizophrenia, carries prognostic information across diagnostic boundaries, as the genetic overlap between the three disorders would predict [56,57]. Bipolar liability did not follow this pattern, although BD and schizophrenia have the highest genetic correlation among the three disorders [111]. The link between liability and outcome may therefore involve components specific to each disorder in addition to shared ones. Genomic factors that cut across diagnoses, estimated by multivariate methods, are the next candidates to test against outcome [57,58].

The effects are small. One SD of MDD-PGS lowered the odds of a good outcome by about 12%, an effect too small to guide the treatment of an individual patient [29,30]. Any clinical value of these scores will come from their combination with clinical information. In GSRD, clinical measures of chronicity and severity classify treatment resistance with good accuracy [14], and models that combined PGS with clinical characteristics improved the prediction of antidepressant efficacy [34] and lithium response [112]. A disorder-liability score is available before the first treatment, does not change over time and indexes a liability that the clinical history captures only indirectly. Pharmacogenomic trials such as PANDORA [73] could include disorder PGS as stratification variables, and larger outcome studies could test whether patients at the top of the MDD-PGS distribution benefit from early augmentation or from treatments with a different mechanism.

### Strengths and limitations

The study has the following strengths: Individual participant data from cohorts with MDD, BD and schizophrenia were analysed with the same model in every cohort. The three scores came from one pipeline, with the same PRS-CS weights from the latest Psychiatric Genomics Consortium GWAS in every cohort, so effect sizes can be compared across scores and diagnoses. The outcome was harmonized from each cohort’s own instrument before any genetic association was computed. The primary analysis was restricted to European genetic ancestry defined from genotype data, with within-cohort principal components and exclusion of duplicates and relatives. Firth regression limited small-sample bias in the smaller cohorts. Sensitivity analyses addressed sample overlap with the discovery GWAS, baseline severity, the outcome definition and the choice of cohorts, and a continuous secondary outcome tested whether the result depended on the binary definition. The study also has limitations. The outcomes came from different instruments and designs: prospective clinical trials (CATIE, STAR*D, PANDORA), naturalistic cohorts (GSRD, Barcelona) and two measures of illness course (STEP-BD, HSRRm). Harmonization to a binary outcome made pooling possible but cannot remove differences in what each cohort measured, and it may have diluted the associations. The main results did not survive FDR correction, and the study had limited power: three cohorts had fewer than 120 patients, and the pooled estimates rest mostly on STAR*D, GSRD and STEP-BD. Six cohorts contributed to the discovery GWAS of their own disorder, and leave-one-out summary statistics were not available. The overlap-free estimate for MDD-PGS had the same size as the primary estimate but a wider interval, which is consistent with a loss of precision. The analysis was restricted to European ancestry, and the results may not transfer to other ancestries [61]. Only 67.7% of SCZ-PGS weight-file variants were scored in the Barcelona cohort. The Barcelona outcome was a retrospective clinical rating with few non-responders, and the HSRRm outcome was a lifetime course rating in which chronic illness without deterioration counted as a good course. Treatment differed between cohorts and was not randomized within most of them, so the analysis cannot separate a prognostic effect from a treatment-specific one. Adherence and dosing were not measured uniformly and could not be adjusted for. Sleep and circadian variables were not harmonized across cohorts and therefore could not be examined as potential mediators or modifiers of the PGS-outcome associations. This may be relevant because sleep disturbance has prognostic significance across these disorders and is also represented in some of the symptom scales used to define treatment response.

### Conclusions

In pooled data from seven cohorts spanning MDD, BD and schizophrenia, a higher polygenic liability to major depression showed a small association with a less favourable treatment outcome. The association had low heterogeneity across cohorts and kept its direction across outcome measures and sensitivity analyses, and schizophrenia liability pointed the same way. Bipolar liability had no overall effect, and its direction depended on the diagnosis. The associations were small and did not survive correction for the three primary tests, and should therefore be considered preliminary. Their consistency across several sensitivity analyses supports further investigation of disorder-liability scores as potential transdiagnostic predictors of treatment outcome. Disorder-liability scores are candidates for multivariable models of treatment outcome, together with clinical predictors, in larger and ancestrally diverse samples.

## Data Availability

Individual-level clinical and genetic data from the NIMH studies (STAR*D, STEP-BD, CATIE) are available from the NIMH Repository and Genomics Resource upon approved application. Data from the European cohorts are available from the respective principal investigators upon reasonable request, within the limits of their ethical approvals. The analysis code and the pre-specified study specification are available from the corresponding author upon reasonable request.

https://www.nimhgenetics.org/

## Acknowledgments

Data were obtained from the NIMH Repository and Genomics Resource (supported by cooperative agreement U24 MH068457), a centralized national biorepository for genetic studies of psychiatric disorders. The NIMH had no role in the design, analysis, interpretation or decision to submit this secondary analysis. The principal investigators and funding sources of each contributing trial are gratefully acknowledged, as detailed below. CATIE: the principal investigators of the Clinical Antipsychotic Trials of Intervention Effectiveness were Jeffrey A. Lieberman, M.D., T. Scott Stroup, M.D., M.P.H., and Joseph P. McEvoy, M.D. The CATIE trial was funded by a grant from the National Institute of Mental Health (N01 MH900001) along with MH074027 (PI: P.F. Sullivan). STAR*D: data and biomaterials were obtained from the limited access datasets distributed from the NIH-supported Sequenced Treatment Alternatives to Relieve Depression (STAR*D) study. STAR*D focused on non-psychotic major depressive disorder in adults seen in outpatient settings, and its primary purpose was to determine which treatments work best if the first medication does not produce an acceptable response. The study was supported by NIMH Contract N01MH90003 to the University of Texas Southwestern Medical Center (ClinicalTrials.gov NCT00021528). STEP-BD: data and biomaterials were collected for the Systematic Treatment Enhancement Program for Bipolar Disorder (STEP-BD), a multi-center, longitudinal project selected from responses to RFP NIMH-98-DS-0001, Treatment for Bipolar Disorder. The project was led by Gary Sachs, M.D., and coordinated by Massachusetts General Hospital in Boston, MA (NIMH grant 2N01MH080001-001).

## Supporting information

**S1 Checklist.** STREGA reporting checklist for genetic association studies.

**S1 Table.** Exploratory all-ancestry estimates, by cohort and pooled.

**S2 Table.** Sensitivity analyses, leave-one-out range and diagnosis subgroups, with the between-subgroup heterogeneity test.

**S3 Table.** Characteristics of all genotyped patients with the primary outcome, regardless of ancestry.

**S4 Table.** Definition of every harmonized variable in every cohort: source variable, coding rule and coverage.

**S5 Table.** Cohort-level associations between each polygenic score and standardized percentage improvement.

**S6 Table.** All coefficients of the primary logistic models of good response or good course, by cohort and polygenic score.

**S7 Table.** All coefficients of the linear models of standardized percentage improvement, by cohort and polygenic score.

